# Predicting Effectiveness of Antihypertensive Medications for Heart Failure based on Longitudinal Patient Records and Deep Learning

**DOI:** 10.1101/2022.11.01.22281804

**Authors:** Shaika Chowdhury, Yongbin Chen, Xiao Ma, Qiying Dai, Yue Yu, Nansu Zong

## Abstract

Drug treatment for heart failure (HF) condition includes different medications. As patients could respond variably to a particular medication, being able to predict drug effectiveness is crucial for personalized treatment. Laboratory tests in EHR summarize different aspects of the patient’s physiological process related to a diagnosis, where blood pressure (BP) is deemed a critical hemodynamic parameter for HF prognosis. This work first proposes a novel method based on combinations of different clinical end points to generate the positive and negative samples corresponding to HF patients on whom the drug is effective and not effective respectively. We then formulate drug effectiveness prediction as a time series classification problem and experiment with several deep learning models, leveraging the temporal BP laboratory measurements from EHR as the features. Over thorough comparative evaluations among 3 categories of HF medications and two types of lab features, we achieved the best F1 performance of ∼0.97.

## 1. Introduction

Heart failure (HF) is a serious cardiovascular disease whereby the heart muscle cannot pump blood sufficiently as per the body’s requirements [1]. With a prevalence of >37.7 million globally [2], HF is characterized by high rates of mortality, hospitalizations and rehospitalizations [3]. Statistically, it is estimated that 35% of HF patients pass away within the first year [1], while 1 in 4 HF patients are readmitted within 30 days of discharge [4].

High blood pressure (BP)/ hypertension is regarded as one of the major risk factors for HF [5]; it can be regulated through drug therapy that lowers BP, known as antihypertensives [6]. There exist different categories of antihypertensives (e.g., ACEI, Beta Blocker, ARB) such that the prescription of a specific medication depends on the symptoms and signs displayed by the patients with HF. For example, ACEI is strongly recommended as an initial therapy for HF patients [7], who are generally symptomless. However, if symptoms such as chest pain or anxiety co-occurs with hypertension then a Beta Blocker medication would be recommended.

Once administered, whether the said medications will be effective on a HF patient is governed by their individual characteristics - genotypes and phenotypes. As a result, a cohort comprising HF patients with similar symptoms might respond either favorably (i.e., *effective*) or unfavorably (i.e., *not effective*) to the same medication with identical dose. Personalizing the treatment trajectories of HF patients using computational approaches would ensure that the pharmacological interventions with the maximally optimized therapeutic effect on each patient are employed.

Most previous studies have performed the task of drug effectiveness prediction as drug response prediction in cell lines [8-12]. These works use the genomic profiles from the cell lines as features to develop predictive models. However, the scarcity of cell line data for many diagnoses poses a major hindrance for pharmacogenomics-based precision medicine.

Electronic health records (EHR) data is a rich source of longitudinal patient information that has recently achieved success in various clinical tasks [13]. As patient’s drug response is solely not dependent on their genomics makeup but also rely on other phenotypic determinants [14], utilizing EHR would facilitate a viable, complementary solution. As a result, some recent works [15, 16] have performed drug response in patients from EHR. A statistical model based on least square formulation is proposed in [15] that models drug response as a linear combination of drug exposures, as well as, time-dependent and time-invariant parameters. While [16] constructs a bipartite graph of drugs and symptoms from EHR and uses a bayesian graphical model. Nevertheless, EHR utility in drug effectiveness prediction is an understudied problem as these are the handful of relevant works to the best of our knowledge.

To address the above limitations, in this work we formulate drug effectiveness prediction as a time series classification problem from EHR using deep learning. The difference between our proposed approach and previous drug response studies using EHR data [15, 16] is twofold. First, modeling EHR as a time series helps to capture the temporal dynamics among the clinical events in the patient’s encounter sequence through the exploitation of the time-varying features. In particular, we leverage the laboratory (lab) tests as the predictive features as they signify numerical values related to various physiological measurements and could potentially provide discriminative signals indicating the course of the HF treatment. For instance, BP lab tests (i.e., DBP, SBP) monitor the overall hemodynamic evolution in HF patients [17], whereupon a relative increase in BP measurement would mean deterioration of patient’s clinical status while a relative decrease in BP value likely reflects the patient’s HF condition is improving. Second, unlike statistical and classical machine learning approaches, deep learning is data-driven and automatically models the complex, non-linear and long-term dependencies in EHR.

Overall, our contributions are summarized as:

- We explore the possibility of using deep learning for drug effectiveness prediction in HF treatment from EHR data. We believe this is the first work that establishes the successful application of deep learning techniques for drug response in patients as time series classification by leveraging the longitudinal phenotypic variables in EHR.
- We propose a novel sampling method based on clinically meaningful endpoints in the treatment of HF, to accurately generate the positive and negative patient instances for better predictive modeling.
- We investigate the effectiveness of three categories of antihypertensive medications in HF treatment using deep learning and achieve best performance of ∼0.97 F1 over a comprehensive evaluation utilizing six neural network models.

## 2. Materials and Methods

### 2.1. Data Source

We use the patient information from the United Data Platform (UDP), the clinical data repository of Mayo Clinic. It is an exhaustive clinical data warehouse that contains millions of patients’ data, which are also updated in real time. It provides a combined view of multiple data sources collected from various clinical and hospital systems within the Mayo Clinic. In this study, we use the time series data recorded in UDP’s EHRs during encounters for the lab tests. Included lab tests are diastolic blood pressure (DBP) and systolic blood pressure (SBP).

### 2.2. EHR Retrieval for HF Patients

To create the appropriate patient cohort and retrieve the relevant EHR data for the study, we undertake three data filtering steps as illustrated in Figure 1. Initially, we identified the patients using diagnosis codes associated with HF, as defined by the ICD-9/ICD-10 codes in Table 1. In this step we also ensure that HF is the patient’s primary medical diagnosis. As a next step, we filtered based on the medication category to ensure that the patient was taking a medication from one of the three antihypertensive categories - Angiotensin-converting-enzyme inhibitors (ACEI), Beta Blocker (BB) and Angiotensin II receptor blockers (ARB) - as listed in Table 2. We chose these three medication categories as they are listed as recommended pharmacological treatment for HF in the 2022 AHA/ACC/HFSA Guideline for the Management of Heart Failure [18]. We further ensure that the HF diagnosis timestamp is earlier than the medication administration timestamp to rule out confounding inclusions. Lastly, as we focus on the use of DBP and SBP lab tests information in this study, so we require the presence of either of the lab measurements in the patient’s EHR. Thereafter, all patients with at least one HF diagnosis code from Table 1, at least one medication from Table 2 and at least two encounters with the DBP/SBP lab tests were included in the corresponding cohort.

**Figure 1.**
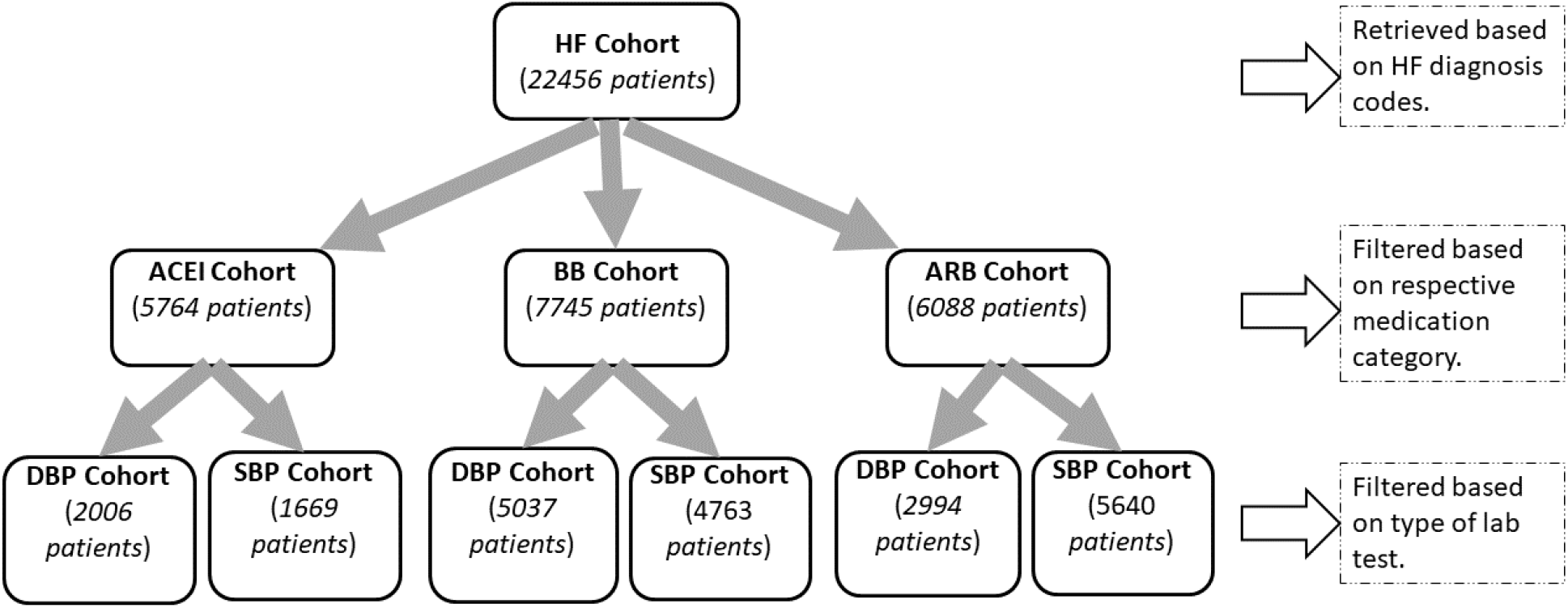
Data Filtering Pipeline. Data size linked to each cohort based on the filtering step is shown within parentheses

**Table 1.** Diagnosis codes associated with HF

|  | ICD Codes |
| --- | --- |
| ICD-9 | 398.91, 402.01, 402.11, 402.91, 404.01, 404.03, 404.11, 404.13, 404.91, 404.93, 428.0, 428.1, 428.20, 428.21, 428.22, 428.23, 428.30, 428.31, 428.32, 428.33, 428.40, 428.41, 428.42, 428.43, 428.9 |
| ICD-10 | I09.81, I11.0, I13.0, I13.2, I50.1, I50.20, I50.21, I50.22, I50.23, I50.30, I50.31, I50.32, I50.33, I50.40, I50.41, I50.42, I50.43, I50.810, I50.811, I50.812, I50.813, I50.814, I50.82, I50.83, I50.84, I50.89, I50.9 |

**Table 2.** Antihypertensive category and medications

| Category | Medication Name |
| --- | --- |
| ACEI | Benazepril, Lotensin, Captopril, Enalapril, Vasotec, Fosinopril, Lisinopril, Prinivil, Zestril, Moexipril, Perindopril, Quinapril, Accupril, Ramipril, Altace, Trandolapril |
| Beta Blocker | Acebutolol, Atenolol, Tenormin, Bisoprolol, Zebeta, Metoprolol, Lopressor, Toprol XL, Nadolol, Corgard, Nebivolol, Bystolic, Propranolol, Inderal, InnoPran XL |
| ARB | Azilsartan, Edarbi, Candesartan, Atacand, Eprosartan, Irbesartan, Avapro, Losartan, Cozaar, Olmesartan, Benicar, Telmisartan, Micardis, Valsartan, Diovan |

### 2.3. Clinical Outcome-based Sampling

As revealed by a seminal work on drug efficacy for clinical trials [19], two types of endpoints are considered clinically meaningful in the evaluation of drugs for HF treatment. The first type evaluates any changes in the patient’s clinical status, while the second evaluates in terms of occurrence of any major clinical event (e.g., death, hospitalization). For the former, New York Heart Association (NYHA) functional classification is used to assess the functional capacity of HF patients based on the severity of symptoms and is considered to provide subjective evidence. Whereas the latter is viewed as a more objective measure as death and hospitalization define definitive changes in the HF progression, thus being less susceptible to observer bias.

Motivated by this work, we design a *composite selection strategy* centered on the second type of measure to separate the positive patients (i.e., the drug is effective) from the negative patients (i.e., the drug is not effective) as EHRs also encompass information related to morbidity and mortality. We also wanted to incorporate the first type of evaluation in our selection criteria, however, based on our early data processing we found that almost all the samples in our cohort belonged to the same NYHA class (i.e., 2-3), which was not distinctive enough for binary class division.

The proposed composite selection strategy is composed of three clinical measures structured as cascaded conditions for more clinically meaningful positive and negative samples, as shown in Figure 2. First, we check for drug withdrawal per category. This is based on the intuition that if the total number of medications for that category (Table 2) in the patient’s encounter history is one, then that could mean that the initially prescribed drug was never replaced with a different drug and hence is a possible indication of drug effectiveness. Nonetheless, reasons for drug withdrawal could also be influenced by the physician’s judgment or other administrative factors [19]. So, we check against two objective measures – mortality and hospitalization - for further validation. If both are false (“No”), then that means the patient did not pass away during the HF treatment route and was not hospitalized for any worsening symptoms, affirming that the drug was effective on the patient. This forms our positive samples. Otherwise, if either of the measures is true (“Yes”) then we assign it as a negative sample. Note that to only count the significant hospitalizations as endpoints, we verify that the minimum duration is 24 hours and the time difference between the current encounter’s admit time and the previous encounter’s discharge time <= 24 hours. Refer to the first condition again for false branching of the number of medications. In this case, there are multiple medications involved in the patient’s encounters so we cannot directly attribute the cause of mortality or hospitalization to the first medication. To mitigate these effects, we only consider the encounters associated with the first medication and analyze the truncated encounter sequence for mortality and rehospitalization. Subsequently, the patient being alive but hospitalized could be traced to the drug not being effective, forming a negative sample.

**Figure 2.**
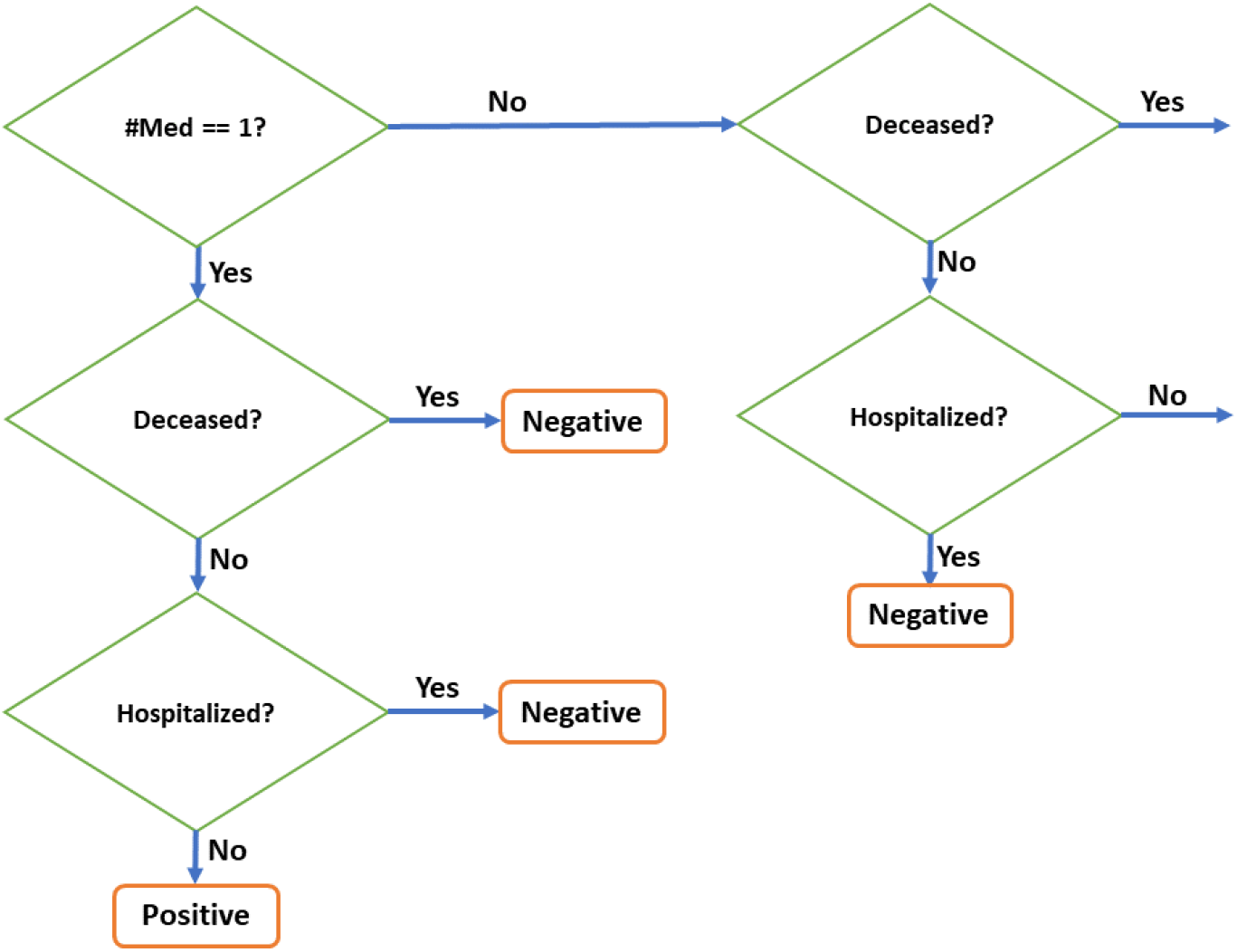
Clinical outcome-based sampling flowchart

### 2.4. Features

In this work, we focus on univariate time series classification, which considers a sequence of time-ordered observations for a single variable as the input. Mapping this to our problem, the patient’s EHR sequence is a list of encounters arranged by the admission time, where the observations within each encounter with respect to the DBP/SBP lab test variable are constructed using the following features.

#### 2.4.1. Lab Measurements

We use the DBP/SBP lab measurements and transform them using Min-Max normalization to scale them in the same range. Table 3 outlines the number of positive, negative and total patient samples in each medication category for the DBP and SBP lab tests using this feature. Note that the positive and negative sample counts are not the same for DBP and SBP as both the lab tests were not taken for some patients.

**Table 3.**
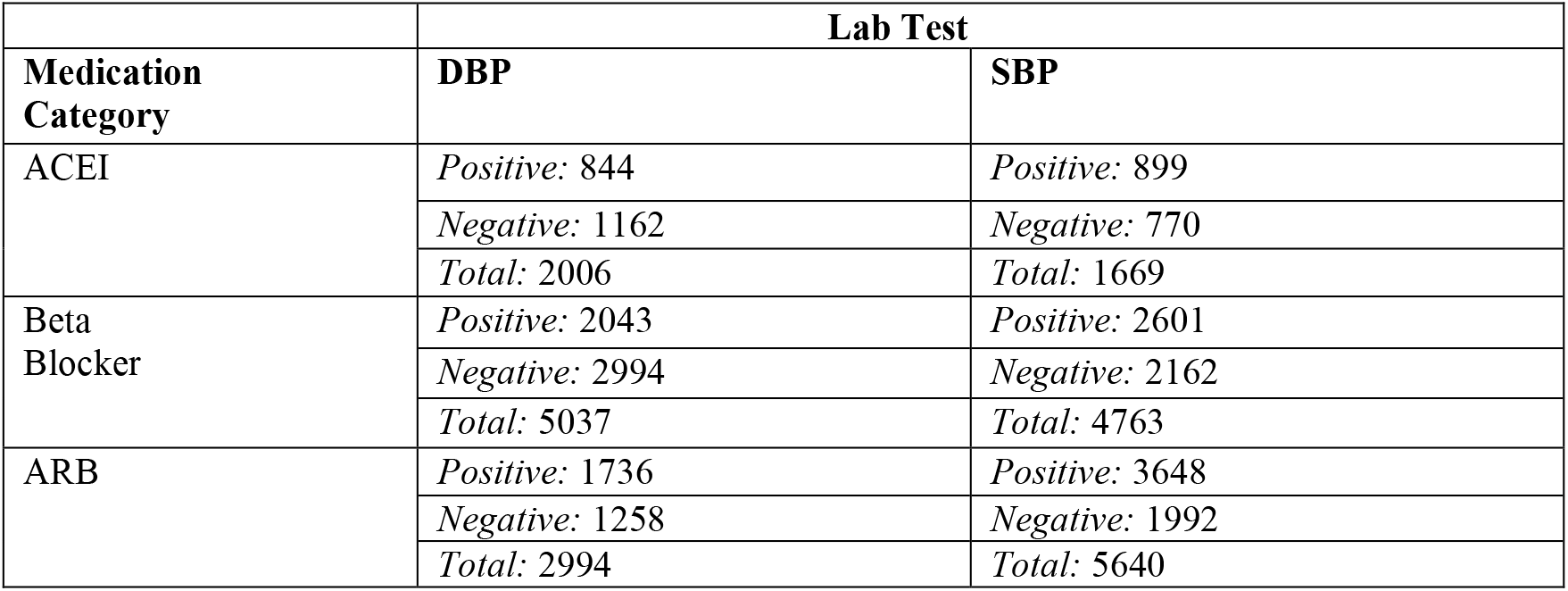
Data statistics for lab measurements feature

#### 2.4.2. Change in Lab Measurements

We define the “duration of effectiveness” as the time period between the drug intake and the time when the first change in DBP/SBP value was observed. That is, for positive samples we consider a decrease in the lab measurement in a subsequent encounter compared to that in the first encounter and for negative samples we consider an increase in lab measurement instead. The intuition is that the decrease/increase in the lab measurement would indicate an improvement/decline in HF status, so we only include the lab measurements in the encounters within the duration of effectiveness. Table 4 outlines the number of positive, negative and total patient samples in each medication category for the DBP and SBP lab tests using this feature. Note that the number of samples for some categories is smaller than its counterpart in Table 3 as now some samples might not have the minimum of two encounters due to the encounter sequence cutoff on the duration of effectiveness, and hence gets excluded from the cohort.

**Table 4.** Data statistics for change in lab measurements feature

| Medication Category | Lab Test |  |
| --- | --- | --- |
|  | DBP | SBP |
| ACEI | *Positive: 380* | *Positive: 144* |
|  | *Negative: 634* | *Negative: 92* |
|  | *Total: 1014* | *Total: 236* |
| Beta Blocker | *Positive: 796* | *Positive: 440* |
|  | *Negative: 1493* | *Negative: 259* |
|  | *Total: 2289* | *Total: 699* |
| ARB | *Positive: 764* | *Positive: 1858* |
|  | *Negative: 608* | *Negative: 1088* |
|  | *Total: 1372* | *Total: 2946* |

### 2.5. Prediction Models

Deep neural networks have exhibited powerful capability in various healthcare [20-23] and biomedical applications [24-27] due to their ability to extract high-level abstract features [28] and learn hierarchical feature representations [29]. To demonstrate the applicability of deep learning techniques in drug effectiveness prediction for HF treatment, we build several neural networks.

#### Multi-layer Perceptron (MLP)

A neural network with one hidden layer of 50 hidden units which is passed through the rectified linear unit (ReLU) activation function [30].

#### LSTM

A variant of recurrent neural network (RNN) [31] that processes the input sequentially by updating the current state based on the past hidden states and current input. Long short-term memory (LSTM) [32] model includes gates in the cells to enable better modeling of long-term dependencies by alleviating the vanishing gradient problem [33]. We set the number of hidden units to 50 and use ReLU.

#### Stacked LSTM (S-LSTM)

This model is composed of 2 LSTM layers stacked together, each with 50 hidden units and ReLU activation. It is a deeper model than vanilla LSTM to enhance prediction performance.

#### Bi-LSTM

A bidirectional LSTM [34] consists of 2 LSTM layers processing the input in opposite directions to facilitate capturing both the previous and future contexts. The hidden unit dimension is 50 with ReLU as the non-linear function.

#### CNN-LSTM

This is a hybrid network that first consists of a convolutional neural network (CNN) [35] component to capture the local information in the input using a 1D convolution and a 1D max-pooling layers. We set the kernel size to 1 and the number of filters to 64. The output is then passed through an LSTM component for the modeling of temporal information in the input. The LSTM layer has 50 hidden units and uses ReLU.

#### Transformer

A non-sequential model, the Transformer [36] uses self-attention mechanism to process the input as a whole, allowing parallel computation. We set the number of attention heads to 1 as a higher number of heads performed worse in our hyperparameter-tuning experiments.

## 3. Results and Discussions

We conducted experiments to investigate the drug effectiveness performance of the deep learning methods on the EHR data and quantified the evaluation using several metrics - Accuracy, AUROC, Precision, Recall and F-1 score. All experiments are executed using 10-fold cross-validation to robustly evaluate the performance of each model. Furthermore, we stratified each fold to ensure the same proportion of positive and negative samples. Each model is trained for 100 epochs in a batch size of 32 using Adam [37] as the optimizer. Implementations of all models are done in TensorFlow v2 [38].

### 3.1. Fine-grained Evaluation based on Facet

We evaluated the deep learning models on three facets - *type of feature, type of lab test* and *medication category* – that examine EHR usefulness in drug effectiveness prediction for HF treatment from different viewpoints for fine-grained analysis. To inspect the contribution of each facet and find the facet combination deriving the best performance, we separately visualize box plots for each facet. When generating the box plots with respect to a facet, the values in relation to the facet form the x-axis, while the other two facets are set to fixed values so as to emphasize the effect that the values of the current facet have on the drug effectiveness performance in a comparative manner. Each box plot shows the distribution of the particular neural network’s performance on the 10-fold test data associated with the facet. We also perform a Student’s t-test to highlight the difference in the performance between the corresponding models through computation of the p-value (ρ). In each figure, we annotate the p-values with placeholder annotations denoting the range the p-value lies in to mark the degree of statistical significance, as described in the legend below. The annotation ‘ns’ stands for not statistically significant difference and star/s indicate statistical significance, such that the higher the number of stars the more statistically significant the difference.

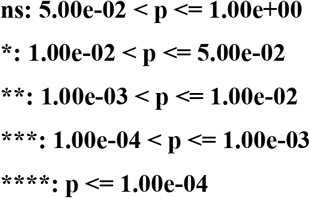

Figure 3 reports the comparison in performance of deep learning models on the facet *feature*. As we are analyzing in terms of the feature, so we hardcode the remaining two facets, lab test and medication category, to the values DBP and ACEI respectively. Thus, this evaluation is carried out to assess the contributions of the two types of DBP features – lab measurement (LM) and change in lab measurement (CLM) – in predicting the effectiveness of ACEI therapy. Comparing the model performances with the LM and CLM features correspondingly in F1 score, it can be seen that the CLM feature consistently outperforms the LM feature for all the models. This is further validated by the p-value as the CLM results are notably statistically significant (p < 0.05) than the LM results across all the models. While the best performing model with CLM feature happens to be S-LSTM. This empirical finding divulges that lab tests are noisy data, possibly due to redundant and erroneous lab values, and considering all the observations (i.e., LM feature) degrades the model’s generalization skill. Instead, only including the measurements that were taken before a drastic change in HF status (i.e., CLM feature) seems to provide informative cues in the model’s learning.

**Figure 3.**
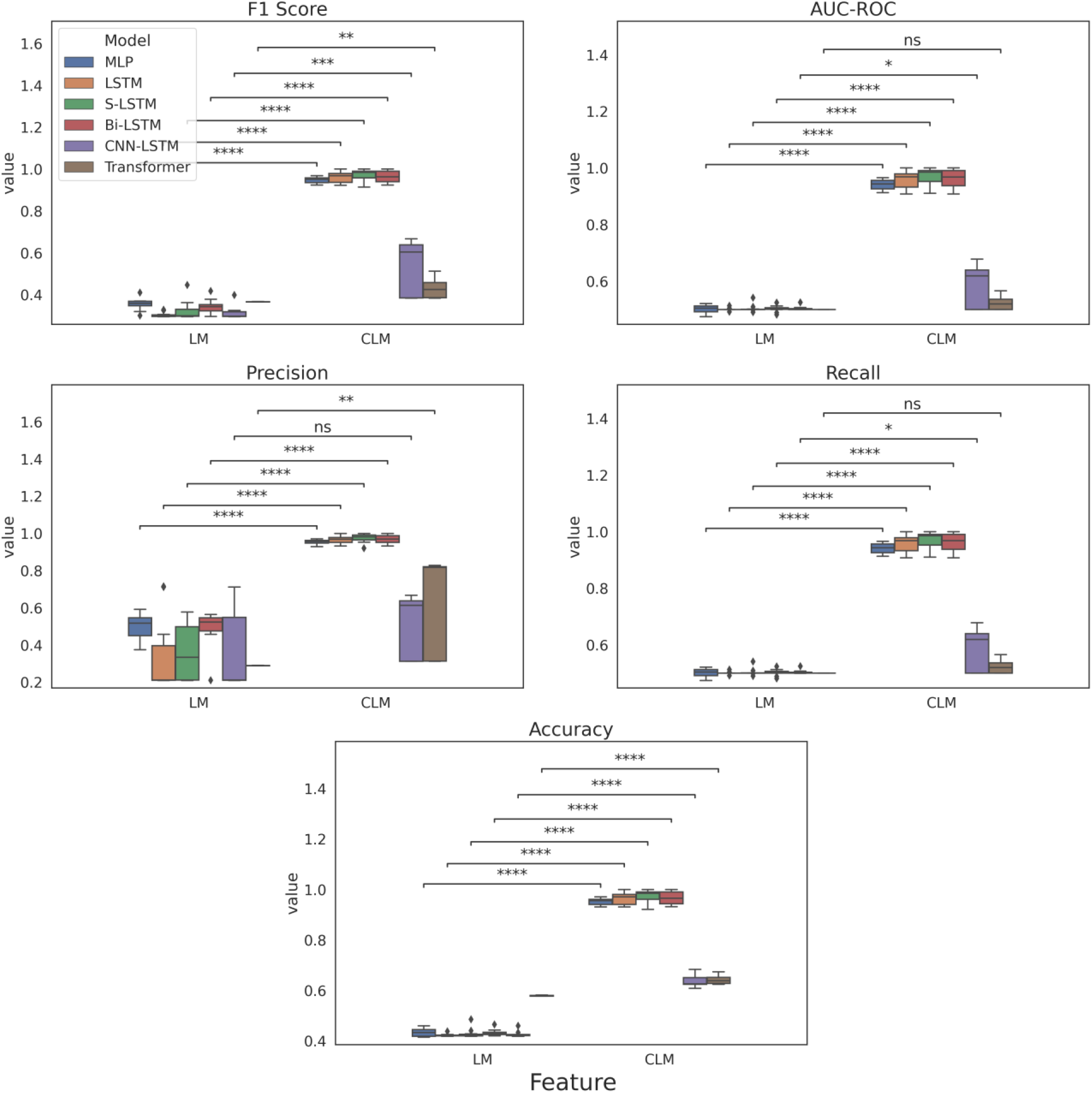
10-fold cross-validation evaluation of deep learning models on the facet *feature*

As DBP and SBP are two main diagnostic lab tests done to track HF prognosis, we then evaluate model performance on the facet *lab test*, presented in Figure 4. In this case, as CLM demonstrated better performance based on the previous evaluation on the facet feature, we fix the value of feature to CLM. While like before, we set the medication category to ACEI. So, this evaluation is performed to elucidate the particular lab test’s impact in predicting the effectiveness of ACEI medications for treating HF. Empirically (i.e., F1 score), from Figure 4 it seems that DBP generally outperforms SBP as five out of the six neural networks exhibit statistical significance, with S-LSTM being the best performing model for DBP.

**Figure 4.**
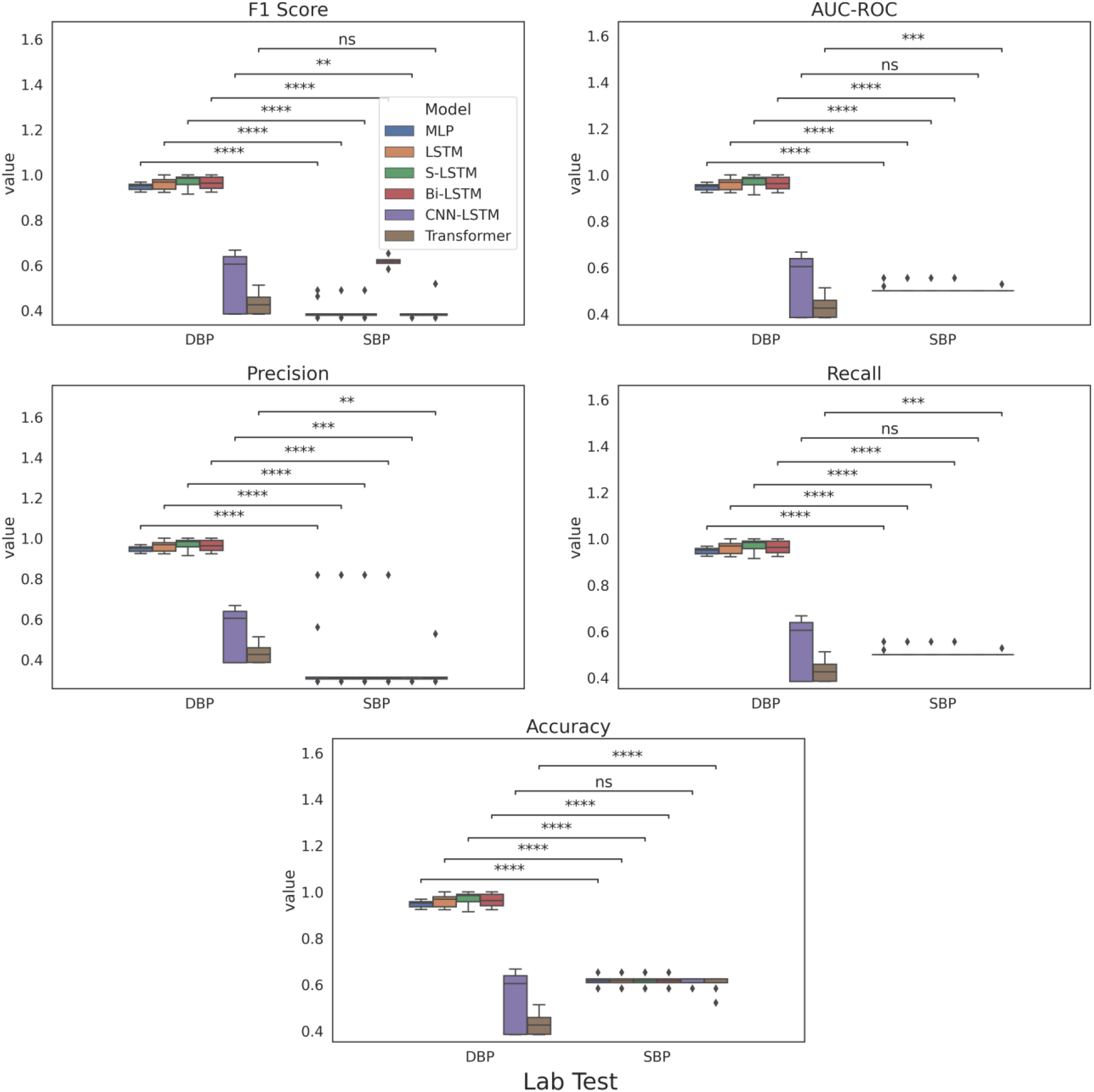
10-fold cross-validation evaluation of deep learning models on the facet *lab test*

Finally, Figure 5 plots the experimental results with the facet *medication category* to reveal the relative performances of the deep models on predicting effectiveness of the three types of antihypertensives – ACEI, Beta Blocker (BB) and ARB. We allocate the remaining facets the best performing values based on the previous evaluations, namely CLM and DBP. Going by the F1 score, on the surface it appears that MLP, LSTM, S-LSTM and Bi-LSTM perform comparably well across all the categories as the F-1 scores for the mentioned models are greater than 0.9. Specifically, S-LSTM performs the best for ACEI, whereas LSTM leads for Beta Blocker and ARB. However, as the differences in performances across the three categories are predominantly not statistically significant, we cannot draw definite conclusions toward the best performing medication category. We assume this finding further corroborates the need for individualized treatment for HF condition that arises from the variability in patients’ responses to different categories of HF medications due to differential patient characteristics. Put another way, this justifies the design of computational models for drug effectiveness using patient-specific data (i.e., EHR) as such.

**Figure 5.**
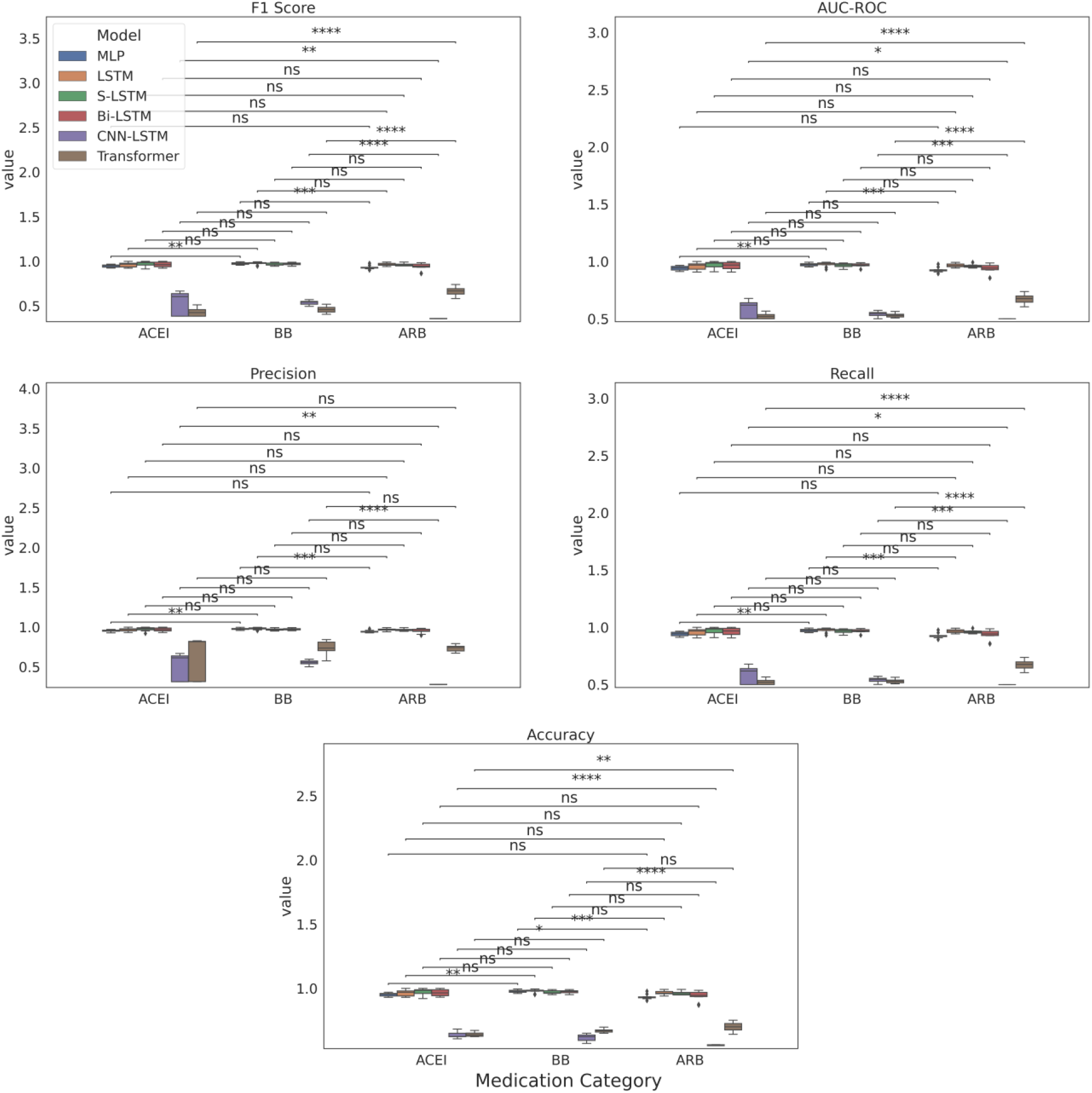
10-fold cross-validation evaluation of deep learning models on the facet *medication category*

The complete tables including all the results covering the two types of features, two types of lab tests, three medication categories and six neural networks could be found in the supplementary section Tables 5-8.

**Table 5.** Performance results per antihypertensive category of deep learning models on DBP time series classification using lab measurements as the feature

|  |  | MLP | LSTM | S-LSTM | Bi-LSTM | CNN-LSTM | Transformer |
| --- | --- | --- | --- | --- | --- | --- | --- |
| ACEI | Accuracy | 0.4339 | 0.4229 | 0.4299 | 0.4329 | 0.4269 | **0.5790** |
|  | AUROC | 0.5011 | 0.5009 | **0.5040** | 0.5027 | 0.5035 | 0.5000 |
|  | Precision | **0.4996** | 0.3356 | 0.3592 | 0.4896 | 0.3624 | 0.2895 |
|  | Recall | 0.5011 | 0.5009 | **0.5039** | 0.5026 | 0.5035 | 0.5000 |
|  | F1-Score | 0.3552 | 0.3032 | 0.3246 | 0.3446 | 0.3129 | **0.3667** |
| Beta Blocker | Accuracy | 0.5073 | 0.5057 | 0.5063 | 0.5069 | 0.5047 | **0.5943** |
|  | AUROC | 0.5138 | **0.5164** | 0.5159 | 0.5162 | 0.5141 | 0.5000 |
|  | Precision | 0.5145 | 0.5172 | 0.5169 | **0.5174** | 0.5151 | 0.2971 |
|  | Recall | 0.5138 | **0.5165** | 0.5159 | 0.5163 | 0.5141 | 0.5000 |
|  | F1-Score | 0.4980 | **0.4995** | 0.4973 | 0.4994 | 0.4977 | 0.3727 |
| ARB | Accuracy | 0.5800 | 0.5800 | 0.5800 | 0.5800 | 0.5800 | 0.5800 |
|  | AUROC | 0.5000 | 0.5000 | 0.5000 | 0.5000 | 0.5000 | 0.5000 |
|  | Precision | 0.2900 | 0.2900 | 0.2900 | 0.2900 | 0.2900 | 0.2900 |
|  | Recall | 0.5000 | 0.5000 | 0.5000 | 0.5000 | 0.5000 | 0.5000 |
|  | F1-Score | 0.3670 | 0.3670 | 0.3670 | 0.3670 | 0.3670 | 0.3670 |

**Table 6.** Performance results per antihypertensive category of deep learning models on DBP time series classification using change in lab measurements as the feature

|  |  | MLP | LSTM | S-LSTM | Bi-LSTM | CNN-LSTM | Transformer |
| --- | --- | --- | --- | --- | --- | --- | --- |
| ACEI | Accuracy | 0.9506 | 0.9634 | **0.9733** | 0.9664 | 0.6377 | 0.6416 |
|  | AUROC | 0.9410 | 0.9586 | **0.9702** | 0.9626 | 0.5838 | 0.5223 |
|  | Precision | 0.9541 | 0.9657 | **0.9740** | 0.9679 | 0.5070 | 0.667 |
|  | Recall | 0.9410 | 0.9586 | **0.9702** | 0.9625 | 0.5839 | 0.5223 |
|  | F1-Score | 0.9465 | 0.9605 | **0.9713** | 0.9638 | 0.5329 | 0.4300 |
| Beta Blocker | Accuracy | 0.9777 | **0.9799** | 0.9707 | 0.9737 | 0.6180 | 0.6700 |
|  | AUROC | 0.9723 | **0.9749** | 0.9667 | 0.9690 | 0.5418 | 0.5293 |
|  | Precision | 0.9786 | **0.9812** | 0.9700 | 0.9737 | 0.5545 | 0.7419 |
|  | Recall | 0.9547 | **0.9748** | 0.9666 | 0.9690 | 0.5417 | 0.5293 |
|  | F1-Score | 0.9752 | **0.9775** | 0.9675 | 0.9709 | 0.5371 | 0.4588 |
| ARB | Accuracy | 0.9343 | **0.9672** | 0.9613 | 0.9416 | 0.5572 | 0.6987 |
|  | AUROC | 0.9271 | **0.9663** | 0.9592 | 0.9358 | 0.5000 | 0.6713 |
|  | Precision | 0.9448 | **0.9680** | 0.9638 | 0.9517 | 0.2786 | 0.7335 |
|  | Recall | 0.9271 | **0.9663** | 0.9592 | 0.9358 | 0.5000 | 0.6713 |
|  | F1-Score | 0.9322 | **0.9667** | 0.9607 | 0.9393 | 0.3578 | 0.6631 |

**Table 7.** Performance results per antihypertensive category of deep learning models on SBP time series classification using lab measurements as the feature

|  |  | MLP | LSTM | S-LSTM | Bi-LSTM | CNN-LSTM | Transformer |
| --- | --- | --- | --- | --- | --- | --- | --- |
| ACEI | Accuracy | 0.5389 | 0.5389 | 0.5389 | 0.5389 | 0.5389 | **0.5467** |
|  | AUROC | 0.5000 | 0.5000 | 0.5000 | 0.5000 | 0.5000 | **0.5131** |
|  | Precision | 0.2694 | 0.2694 | 0.2694 | 0.2694 | 0.2694 | **0.4106** |
|  | Recall | 0.5000 | 0.5000 | 0.5000 | 0.5000 | 0.5000 | **0.5131** |
|  | F1-Score | 0.3502 | 0.3502 | 0.3502 | 0.3502 | 0.3502 | **0.4444** |
| Beta Blocker | Accuracy | **0.5461** | **0.5461** | **0.5461** | **0.5461** | **0.5461** | 0.5426 |
|  | AUROC | **0.5000** | **0.5000** | **0.5000** | **0.5000** | **0.5000** | 0.4979 |
|  | Precision | 0.2730 | 0.2730 | 0.2730 | 0.2730 | 0.2730 | **0.2919** |
|  | Recall | **0.5000** | **0.5000** | **0.5000** | **0.5000** | **0.5000** | 0.4979 |
|  | F1-Score | 0.3532 | 0.3532 | 0.3532 | 0.3532 | 0.3532 | **0.3649** |
| ARB | Accuracy | **0.6469** | **0.6469** | **0.6469** | **0.6469** | **0.6469** | 0.6446 |
|  | AUROC | **0.5000** | **0.5000** | **0.5000** | **0.5000** | **0.5000** | 0.4993 |
|  | Precision | 0.3234 | 0.3234 | 0.3234 | 0.3234 | 0.3234 | **0.4333** |
|  | Recall | **0.5000** | **0.5000** | **0.5000** | **0.5000** | **0.5000** | 0.4993 |
|  | F1-Score | 0.3928 | 0.3928 | 0.3928 | 0.3928 | 0.3928 | **0.4522** |

**Table 8.** Performance results per antihypertensive category of deep learning models on SBP time series classification using change in lab measurements as the feature

|  |  | MLP | LSTM | S-LSTM | Bi-LSTM | CNN-LSTM | Transformer |
| --- | --- | --- | --- | --- | --- | --- | --- |
| ACEI | Accuracy | **0.6144** | **0.6144** | **0.6144** | **0.6144** | 0.6126 | 0.6039 |
|  | AUROC | **0.5073** | 0.5054 | 0.5054 | 0.5054 | 0.5000 | 0.5027 |
|  | Precision | **0.3819** | 0.3564 | 0.3564 | 0.3564 | 0.3063 | 0.3285 |
|  | Recall | **0.5075** | 0.5055 | 0.5055 | 0.5055 | 0.5000 | 0.5027 |
|  | F1-Score | **0.3983** | 0.3899 | 0.3899 | 0.3899 | 0.3798 | 0.3938 |
| Beta Blocker | Accuracy | 0.6280 | 0.6309 | 0.6280 | **0.6480** | 0.6303 | 0.6303 |
|  | AUROC | 0.4996 | 0.5035 | 0.4988 | **0.5274** | 0.5000 | 0.5000 |
|  | Precision | 0.3646 | 0.3989 | 0.3144 | **0.4624** | 0.3151 | 0.3151 |
|  | Recall | 0.4997 | 0.5034 | 0.4988 | **0.5273** | 0.5000 | 0.5000 |
|  | F1-Score | 0.3894 | 0.3968 | 0.3857 | **0.4338** | 0.3866 | 0.3866 |
| ARB | Accuracy | **0.8669** | 0.6306 | 0.6306 | 0.6710 | 0.6309 | 0.6512 |
|  | AUROC | **0.8329** | 0.5000 | 0.5000 | 0.5594 | 0.5000 | 0.5497 |
|  | Precision | **0.8833** | 0.3153 | 0.3153 | 0.5800 | 0.3154 | 0.6350 |
|  | Recall | **0.8329** | 0.5000 | 0.5000 | 0.5594 | 0.5000 | 0.5497 |
|  | F1-Score | **0.8476** | 0.3867 | 0.3867 | 0.4974 | 0.3868 | 0.5134 |

### 3.2. Varying the Time Series Length

In the practical healthcare setting, the length of the time series data in EHR could be unequal as patients could have different number of encounters representing their treatment journey. To mimic this real-world trend for more reliable prediction of drug effectiveness, we henceforth evaluate the deep learning models by varying the time series length (i.e., maximum number of encounters). For this experiment, we set the three facets to the values CLM, DBP and ACEI as a case study and evaluate using the best performing model on this cohort from the previous evaluations, namely S-LSTM. On average, patients have 2 encounters in this cohort; so we vary the sequence length of the time series to the following values - 2, 3, 4 and 5 - which results in datasets of sizes 750, 145, 65 and 31 patients respectively. The 10-fold cross-validation performance of the S-LSTM model on this cohort with time series data of different lengths as the input is shown in Figure 6. The best performance was achieved by the time series of length 3, although this performance is comparable to that with length 2 as there is no statistical significance between them. While on increasing the time series length further, the performance degrades. We assume this degradation in performance could be due to the fewer samples available for training with lengths 4 and 5, which caused the model to overfit.

**Figure 6.**
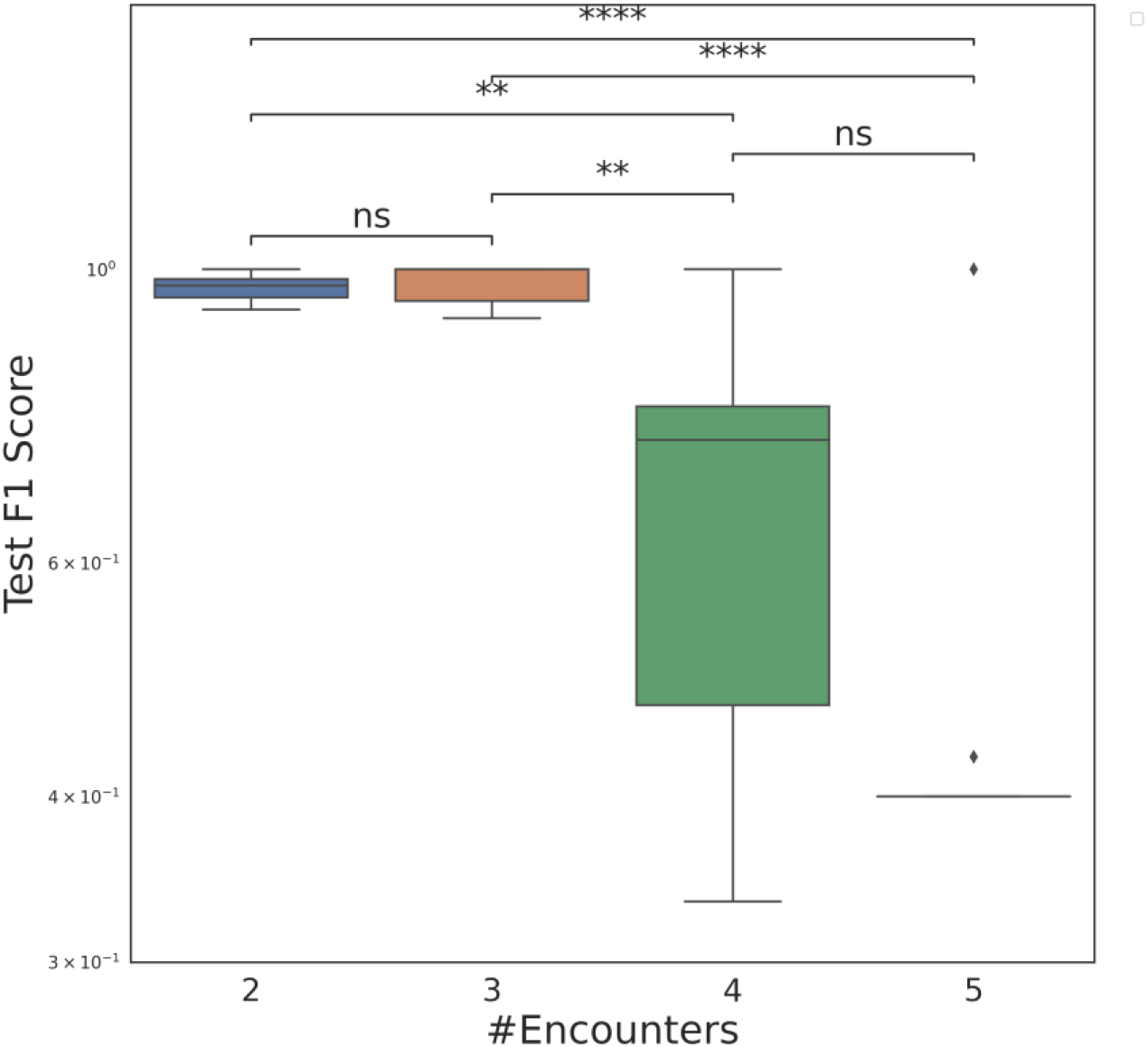
drug effectiveness prediction with different time series lengths

## 4. Conclusions and Future Directions

A predictive model that can help with the identification of drug effect on patients has the scope to support clinical decisions that would enable more accurate prognosis and timely intervention for HF treatment. Towards this goal, in this work we show that deep learning based predictive models for drug effectiveness prediction assert promising results using the lab test data in EHR. We propose a sampling method that comprehensively checks against medically relevant measures to precisely differentiate between cases of drug effectiveness and ineffectiveness pertaining to HF treatment. Extensive experiments covering three categories of antihypertensive medications, two types of lab tests and features, and six deep learning models have been performed.

We acknowledge that there are some limitations of this work that we would like to probe as future directions. In the current study, univariate time series classification models the DBP time series independent of the SBP time series and vice versa. In future work, it would be interesting to account for the interactions between the DBP and SBP clinical variables via a multi-variate time series formulation. In this study, we only use one data source in EHR (i.e., lab tests); while this eases the complexity of time series input modeling as lab measurements are inherently a continuous variable and also helps to directly embody information related to the patient’s physiological process conveniently, it falls short of capturing the heterogeneity in EHR as it fails to incorporate the phenotypic variables present in the other data modalities in EHR (e.g., notes). Integrating the physiological time series data with clinical notes and modeling jointly would open avenue to future investigations in drug effectiveness prediction.

## Data Availability

All data used in the present study are PHI so cannot be make publicly available.

## Acknowledgements

This study is supported by the National Institute of Health (NIH) NIGMS (R00GM135488).

## Supplementary Materials

In each table, the best performance result for each evaluation metric is formatted in bold.

